# Improving experience of medical abortion at home in a changing therapeutic, technological and regulatory landscape: a realist review

**DOI:** 10.1101/2022.07.13.22277600

**Authors:** Paula Baraitser, Caroline Free, Wendy Norman, Maria Lewandowska, Rebecca Meiksin, Melissa Palmer, Rachel Scott, Rebecca French, Kaye Wellings, Alice Ivory, Geoffrey Wong, The SACHA study team

**Author notes:** Corresponding author: Maria Lewandowska, 020 7927 2761, London School of Hygiene and Tropical Medicine 15-17 Tavistock Place, WC1H 9SH.

## Abstract

**Objectives:**

- To synthesise evidence on user experience of medical abortion at home
- To develop a realist programme theory to explain what interventions improve user experience for whom and in what context.
- To use this programme theory to develop recommendations for service providers and those having medical abortions at home

**Background:** Changes in the therapeutic, technological and regulatory landscape are increasing access to medical abortion at home. This intervention is safe, effective and acceptable to most. Clinical pathways and user experience are nevertheless variable and a minority would not choose this method again. We synthesised evidence to inform service development and responsiveness for different people and contexts.

**Methods:** We used a realist approach to literature review that starts from an initial programme theory and generates causal explanations in the form of context-mechanism-outcome configurations to test and develop that theory. We searched the literature 01/01/2000 - 09/12/2022 using broad search terms and then selected papers for their relevance to theory development in contexts relevant to service development in the UJ. Data were analysed using a realist approach to analysis to develop causal explanations.

**Results:** Our searches identified 12,517 potentially relevant abstracts with 835 selected for the full text assessment and 49 papers included in the final review. Our synthesis suggests that having a choice of abortion location remains essential as some people are unable to have a medical abortion at home. Choice of place of abortion (home or clinical setting) was influenced by service factors (number, timing and wait for appointments), personal responsibilities (caring/work commitments), geography (travel time/distance), relationships (need for secrecy) and wish to be aware of/involved in the process. We found that the option for self-referral through a telemedicine consultation, realistic information on range of experiences, opportunities to personalise the process, improved pain relief and choice of when and how to discuss contraception could improve experience.

**Discussion:** Acknowledging the work done by patients when moving an intervention from clinic to home is important. This includes preparing a space, managing privacy, managing work/caring obligations, deciding when/how to take medications, understanding what is normal, assessing experience and deciding when and how to ask for help. Strategies that reduce surprise or anxiety and enable preparation and a sense of control support the transition of this complex intervention outside healthcare environments.

**Strenghts and limitations:**

– Strengths: systematic and transparent approach to the realist review, which was conducted in accordance with the RAMSES standards (27); Authorship team represents a variety of clinical and academic backgrounds
– Limitations: analysis on publicly accessible literature, located through recognised research databases and Google; there were gaps in the evidence that we found and we have highlighted these in our conclusions.

## Background

Changes in the therapeutic, technological and regulatory landscape are impacting abortion care and changing access to medical abortion at home. Medical abortion requires two medications, mifepristone (a competitive progestogen receptor antagonist) and the synthetic prostaglandin, misoprostol. Since mifepristone was first licenced for use in Great Britain in 1991, the proportion of all abortions that are medical abortions has increased steadily with a more recent shift towards medical abortion at home, where both the mifepristone and misoprostol or the misoprostol alone are taken at home. During the COVID 19 pandemic these trends were accelerated and in 2020, 85% of all abortions completed in England and Wales, and 88% of those in Scotland, were medical abortions at home^1,2^. These developments reflect similar changes in Australia, Canada and Europe^3-7^.

Medical abortion at home is safe, effective and acceptable and clinical guidance on some elements, such as doses of abortion medications, are consistent^7-21^. However, significant variation in other elements of clinical pathways remain as providers test new approaches to providing information, support, pain relief and follow up^22^. It is important to note that a minority (17 – 34%) of people who participate in research on medical abortion at home would not choose this method if having an abortion again^12,23-25.^ We are interested in the evidence to inform optimal configuration of medical abortion services at home and particularly in the differences in experience between populations and contexts.

### Terminology and settings

Our findings relate to settings where abortion is legal and where there is a choice of abortion method and setting. Recommendations may be different in settings where abortion is illegal, where mifepristone is not available, where medication combinations or doses are used that are nt evidence based or where surgical options are not available or not safe. Women and people with other gender identities may have abortions. We have used the terms ‘people who have an abortion’ or ‘people’ to describe our population of interest.

### Review question

- What support, for whom, and in what circumstances could improve the experience of people who have medical abortion at home?

## Methods

The SACHA study brings together over 20 researchers from seven countries to create an evidence base to inform health service configuration for abortion provision in the UK (https://www.lshtm.ac.uk/research/centres-projects-groups/sacha#welcome). This review is part of this programme of work and draws on the expertise of the team.

We conceptualised medical abortion as a complex process that has context-sensitive outcomes which may be variably delivered and experienced. A realist approach enabled us to both account for and explain the influence of context on this variation^26^. Our protocol was registered with Prospero https://www.crd.york.ac.uk/prospero/display_record.php?RecordID=225307 and we followed the steps which are commonly used in realist reviews^27,28^-location of existing theories, evidence search and document selection, data extraction, data synthesis and development of a refined programme theory.

We used the PRISMA checklist when writing our report.^1^

### Location of existing theories

A realist approach to understanding interventions (in our case home medical abortion) proposes that any intervention is underpinned by one or more theories (that may be implicit or explicit) in the design of the intervention. This theory sets out how the designers anticipate their intervention will work. In a realist review this understanding is captured via an initial programme theory. Our initial programme theory was developed with input from the SACHA team. We charted the stages of home medical abortion from a user perspective to understand what type of support might be required from the health care system at each stage (figure 1). These stages within a user journey were used to structure our analysis.

**Figure 1 :**
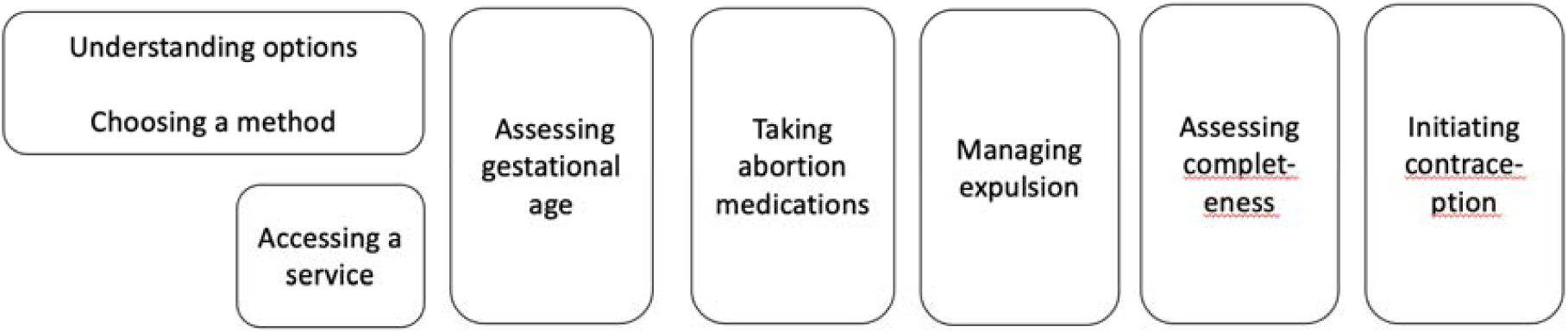
Stages of the medical abortion process (initial theory of change)

### Searching for evidence and document selection

Our evidence search strategy started with a comprehensive literature search designed to answer a broader question for the SACHA project – this was a scoping review that sought to address the question: ‘Interventions of models of abortion care/provision: addressing current therapeutic, technological and regulatory trends which would be relevant to or feasible in the UK in the next 5 years.’ The databases and search terms for this search are listed in appendix 1 and the inclusion and exclusion criteria are listed in figure 1. The search dates were 01/01/2000 - 09/12/2022.

**Figure 1:**
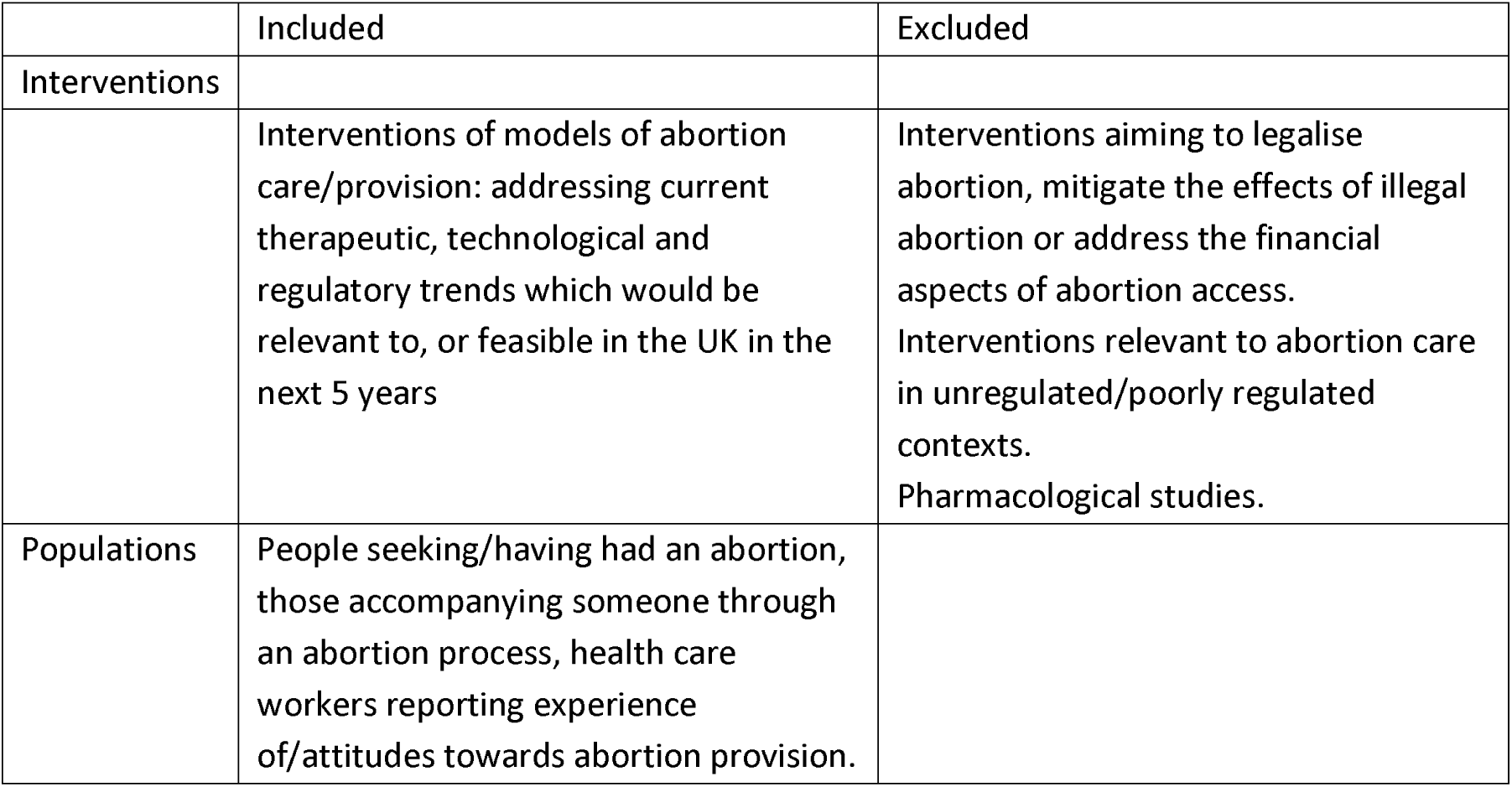
Inclusion/exclusion criteria for scoping review.

Search results were imported into an electronic reference manager Covidence (https://www.covidence.org/) and screened in two stages. The first screen was based on title and abstract with a sample double-screened to check for consistency and discussion of studies of uncertain eligibility to reach consensus. The remainder of papers at the abstract and title stage and all those at the full text stage were single screened. The full text of all included documents were uploaded into a second electronic reference management system (Endnote).

We were confident that this search was comprehensive and contained all relevant information that was needed to answer our realist review question. We subsequently re-screened this search for the purposes of the realist review as follows:

1. A subject expert (WN) screened all full texts to identify documents that were specifically related to medical abortion at home and then grouped these into the stages outlined in our intitial programme theory.
2. PB read these papers to identify whether they were relevant to developing our initial programme theory.

Papers were considered useful for theory development and testing where they offered empirical evidence that could refine, refute or confirm developing emerging realist programme theory and inform the development of context-mechanism-outcome (CMO) configurations within it^28^. This meant including all papers that were relevant to people’s experience of each stage of the medical abortion at home journey and the support provided that might modify this experience. We used a broad definition of ‘support’ to include any intervention by the individual having the abortion, the health care system or friends and family intended to improve or maintain health or wellbeing before, during and after the abortion process.

Realist review processes are iterative so as our CMO configurations developed we revisited the list of included papers for the scoping review to look for additional relevant materials. We also completed additional searches as required including handsearching reference lists and completing new searches, to seek out more relevant data.

### Data extraction

The final selected papers regarding each stage of abortion care were read and re-read by PB and, for the post abortion contraception, by CF. Findings were summarised in spreadsheets which contained information on key relevant findings from each paper, grouped according to the programme theory. Based on their interpretations of these findings PB and CF developed CMO configurations (CMOCs) for each stage of the medical abortion process. No uniform data set was extracted from each paper, rather the data (verbatim sections of text) of each paper that were relevant to each emerging CMOCs were grouped together and iteratively used during the analysis process (see below) to develop CMO configurations.

### Data analysis and synthesis

A realist logic of analysis uses data to produce causal explanations for outcomes that occur within a programme theory in the form of CMOCs. A CMOC is a proposition that explains what element of an initiative works, for whom and in what circumstances and is the primary way of reporting findings within a realist review. Within a CMOC, the causal claim being made is (in its simplest form), when a particular context is present, it ‘triggers’ or ‘activates’ a particular mechanism, which causes a particular outcome. Mechanisms are hidden causal processes that are context-sensitive and are usually inferred based on interpretations of the data. Data to inform our interpretation of the relationships between contexts, mechanisms and outcomes were sought within and across documents (e.g. mechanisms inferred from one document helped explain the way contexts influenced outcomes in a different document). Synthesising data from different documents is often necessary to compile CMOCs, since not all parts of the configurations will be articulated in every document. During our analysis, we used interpretive cross-case comparison to understand and explain how and why reported outcomes have occurred.

### Refining programme theory

We iteratively refined our initial programme theory as the review progressed based on our interpretations of the data from the included papers. For each stage of the abortion at home process theory we sought to unpack what support is needed. Our final programme theory contains CMOCs that explain the outcomes achieved by the support provided, why it happens and in which contexts.

## Results

Our searches identified 27,982 potentially relevant abstracts for both reviews. After duplicates and papers with incorrect dates or excluded literature types were removed 12,401 titles and abstracts were screened with 944 selected for full text assessment. At this stage, 590 studies were excluded which did not meet the inclusion criteria or were duplicates. The remaining full texts were screened for potential relevance to the realist review question and 244 studies were categorised by a content expert (WN) against the inclusion criteria and the initial programme theory. Additional detailed screening of these 244 studies indicated that 35 contained data that were relevant to support theory building and testing and hence were included. A further 12 papers were identified as relevant during the analysis through hand searching and expert consultation. At the end of the analysis, updating the searches identified 43 new abstracts of which 3 were included giving a total of 50 included papers (Figure 3). A table summarising the included papers is given in appendix 2.

**Figure 3:**
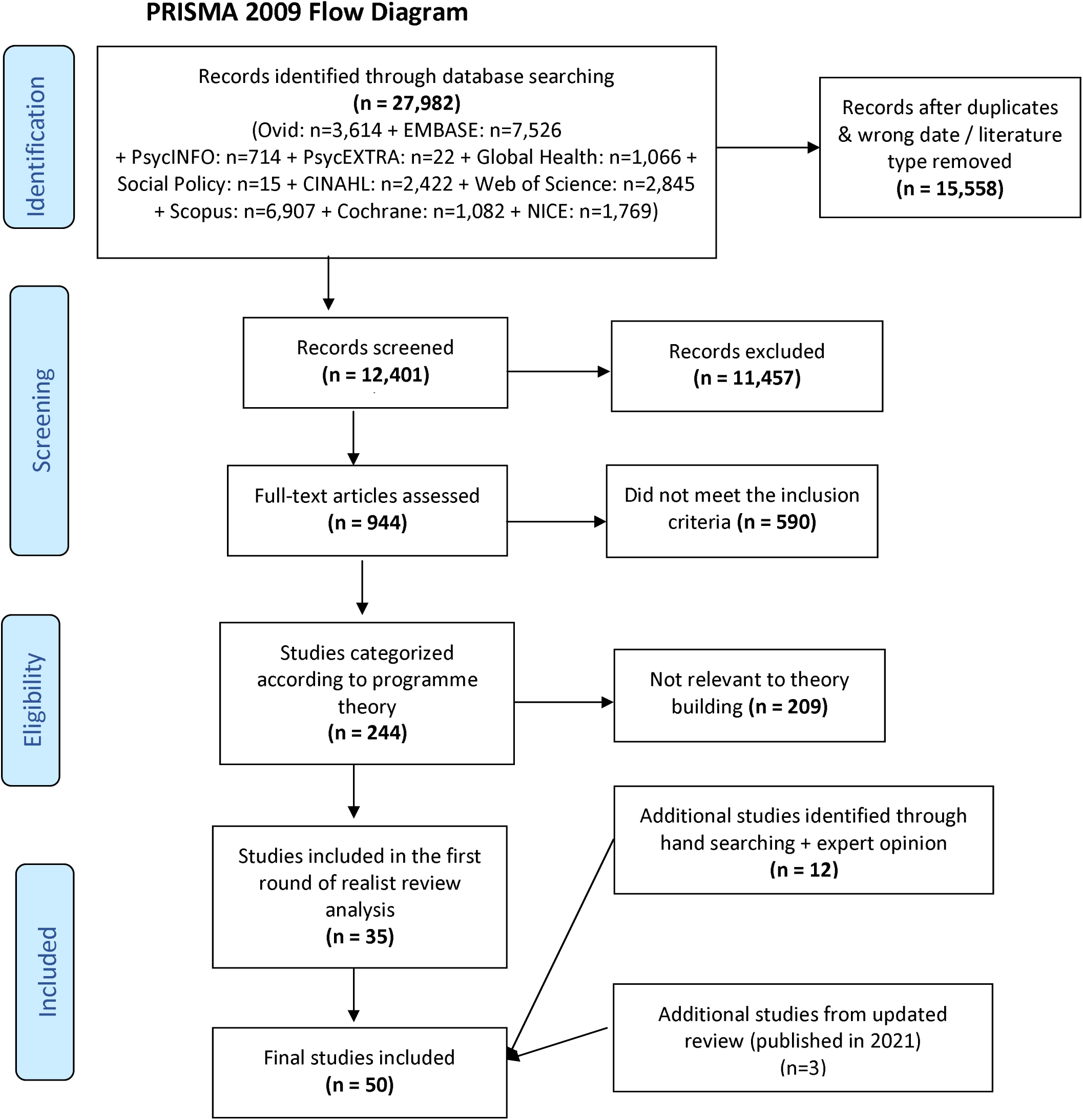
PRISMA diagram

Our results are structured by our programme theory. For each stage, we present a narrative explanation of our findings followed by a table of the CMOCs that underpin the explanation. We found that the evidence on some of the stages in our programme theory were clustered and therefore the following stages are presented together:

- ‘Understanding abortion options’ and ‘Choosing a method/setting’
- ‘Accessing an abortion’ and ‘Pre-abortion counselling assessing gestational age’
- ‘Taking mifepristone’ and ‘Taking misoprostol’.

### Understanding abortion options and choosing a method

Choice of abortion methods and settings was influenced by service factors (number of appointments, timing and wait for appointments), personal responsibilities (caring/work commitments), geography (travel time/distance), relationships and need for privacy/secrecy and the extent to which people wished to be aware of/involved in/in control of the process ^12, 29-35^(Table 1). Those choosing medical abortion at home made this decision on the basis of their social responsibilities (such as the need to provide childcare) or when a home abortion enabled them to maintain their confidentiality, rather than their personal preferences about the procedure itself^12,21,29,35,36,37^. Conversely, some people chose hospital care because it limits knowledge of the procedure among household members (for example, an abusive or unsupportive partner) and requires less responsibility for the process^32,34^. Although we were not specifically seeking literature on reasons for surgical abortion, people cited a wish to be asleep/unaware of the procedure or to avoid pain/bleeding/sight of the products of conception ortaking the mifepristone^36,38^ or because it could be completed quickly^39^.

**Table 1:** CMOCs for ‘understanding options’ and ‘choosing a method’

|  | CMOC |
| --- | --- |
| 1 | When people who want an abortion are offered a choice of procedures (c) they are more likely to be satisfied with their experience (i.e. predict choice of this method again) (o) because they feel their decision is their own (i.e. empowered in their choice) (m).<sup>24, 29, 36, 37, 38, 40, 44, 45</sup> |
| 2 | Where the information provided about each procedure option includes clinical safety and effectiveness and experiential information (c) people are in a better position to make an informed choice (o) because they are more likely to have the information needed to do so (m)<sup>41, 46</sup>. |
| 3 | Where a range of personal experience is included that includes realistic accounts of pain/bleeding (c) then people are more likely to be satisfied with their choice (o) because they |
|  | know what to expect and feel in control of the process (m)<sup>31,39-43</sup> |
| 4 | When people have social responsibilities in their lives (c), they will choose the method for abortion that fits best with these responsibilities (o) because of convenience (m)<sup>21,29, 31,48</sup> |
| 5 | When people would prefer not to take an active role in the abortion process (c), they may choose a surgical abortion (o) because it enables them to distance themselves from the abortion process (m)<sup>23,31,36,47,48</sup> |
| 6 | When people have doubts about their abilities to carry out a medical abortion properly (c), they are more likely to request they are closely supervised (e.g. complete the abortion within a health care context) (o), because of the emotional and practical support provided (m)<sup>36-38,</sup> |
| 7 | When people have had what they consider to be an unpleasant experience of medical abortion (c), they will be unlikely to choose medical abortion again (o), because they want to avoid the experience (m)<sup>24,38,39,37,42,</sup> |

People valued frank descriptions from health professionals about the range of experience of medical abortion^40,31,39-43^. A participant in one study commented that ‘It’s good to do it at home but I think a good level of honesty from the nurse is helpful because it’s on the cusp of being copeable with’^38^. Fear about the abortion procedure was the most frequently cited barrier to accessing an abortion in one study, with information as the most highly ranked facilitator to care^41^. High levels of satisfaction were associated with information on abortion delivered through short films in clinical contexts ^43-45^. It is notable that in this study the film was rated as more informative than a leaflet even though the content was the same. Developing high quality information resources is challenging because of the need to offer both clear clinical instructions and a wide range of experience that enables people to prepare themselves, their supporters and their surroundings. One study evaluating an animated account of one person’s experience of medical abortion at home showed that this approach was well received but participants were concerned that their own experience was different from that shown^43^.

### Accessing a service/pre-abortion consultation/assessing gestational age

People value telephone/telemedicine self-referral options^10,12,21^, because of the privacy, autonomy and convenience they potentially afford and because they remove risk of judgmental attitudes from referring professionals (Table 2). People rated distance to clinic and waiting times as more important than whether counselling was via telemedicine or face to face^30^ and reasons for preferring a telemedicine consultation included the ability to engage from a more comfortable space (at home) and feeling more able to ask questions^49^. However, some people need to take action to speak privately, for example, completing the consultation in their car^49,50^.

**Table 2:**
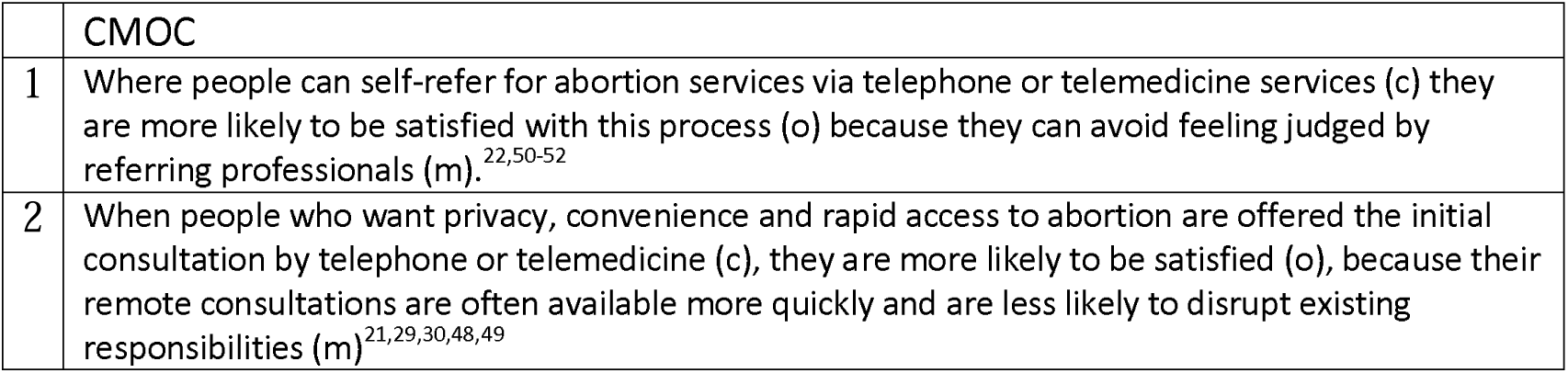
CMOCs for <u>‘Accessing a service/pre-abortion consultation</u>

|  | CMOC |
| --- | --- |
| 1 | Where people can self-refer for abortion services via telephone or telemedicine services (c) they are more likely to be satisfied with this process (o) because they can avoid feeling judged by referring professionals (m).<sup>22,50-52</sup> |
| 2 | When people who want privacy, convenience and rapid access to abortion are offered the initial consultation by telephone or telemedicine (c), they are more likely to be satisfied (o), because their remote consultations are often available more quickly and are less likely to disrupt existing responsibilities (m)<sup>21, 29,30,48,49</sup> |

The pre-abortion consultation is variable and may include an initial in-person consultation with/without an ultrasound scan and/or an examination or a fully remote service including self-reporting of gestational age^22^. Medical abortion without a prior ultrasound scan is safe^12^ and in one qualitative study showed that some people preferred not to have an ultrasound scan because they associated it with having positive experiences of desired pregnancy^49^ and in another study ultrasound was rated as unimportant for 53% of participants.

### Taking the abortion medications

Most of the literature focuses on the experience of taking misoprostol because this generates the most physical side effects of medical abortion; however, some studies suggest that people may find taking the mifepristone the most emotional part of the treatment regimen and that this should be acknowledged in the information and support provided^40^ (table 3). Self-administering misoprostol enabled people to control the timing of their abortion and prepare by making a comfortable space or obtaining things that they might need such as analgesia or hot water bottles, fit the abortion around other responsibilities or keep it private^24^. When given a choice of how to take the misoprostol, 70% in one study chose to self-administer it sublingually, rather than vaginally because they felt more confident that they had inserted it correctly using this approach^21^. Side effects from the misoprostol were common, with 77% of people experiencing one of the common symptoms (nausea, vomiting, diarrhoea or headaches)^21^.

**Table 3:** CMOCs for ‘Taking the abortion medication’

|  | **CMOC** |
| --- | --- |
| 1 | When people who want an abortion have reservations about their active participation in the process (c) they may prefer not to self-administer the medications (o) because they feel personally responsible for ending the pregnancy (m)<sup>40</sup> |
| 2 | When people are given the option of self-administering misoprostol (c) they are better able to prepare for the process (o) because they can choose the time and place of the medical abortion (m)<sup>24</sup> |
| 3 | When people lack confidence in their ability to self-administer misoprostol (c) they prefer that this be done by a health professional (o) because they can then be confident that the procedure is done correctly (m)<sup>21,24,38</sup> |

### Managing expulsion

Experience of managing the expulsion of the products of conception is highly variable (Table 4). People interviewed after medical abortion at home point to the importance of understanding ‘what is normal’ as an important factor to help manage anxiety and to guide seeking clinical advice. Remote clinical support is important and highly valued and is used by 10-50% of people who choose medical abortion at home ^24,37,53,21^.

**Table 4:** CMOCs for ‘Managing expulsion’

|  | CMOC |
| --- | --- |
| 1 | Where people experience pain or bleeding that is outside the limits of what they were told to expect (c) they are more likely to request clinical support (o) because they are concerned that their health may be at risk (m) <sup>23,24,37</sup> |
| 2 | Where people having medical abortion at home have also had a previous pregnancy (live birth/abortion/miscarriage) (c), they are less likely to report pain that is distressing and to request professional support (o) because the experience is more familiar (m) <sup>37,50</sup> |
| 3 | Where people receive adequate pain relief during medical abortion (c) they are more likely to be satisfied with the whole experience of medical abortion at home (o) as they were more comfortable (m) <sup>24,37,38</sup> |
| 4 | Where people want to have privacy when having an abortion (c), they may choose to manage the abortion process at home (o) because they feel less exposed (m) <sup>35,38</sup> |
| 5 | Where people have family/friends/partners whom they know will be supportive when they have an abortion (c) then they value their presence during a medical abortion at home (o) because of the practical and emotional help that they provide (m). <sup>24,56</sup> |

While most people find the process acceptable, the evidence shows that some experience pain and bleeding that is unexpected or distressing and cite this as the reason for future preference for an alternative method ^24,37,38^. In one study 94% of respondents had experienced pain with a median reported pain score of 6.7 (on a scale of 1-10) though 57% reported their pain as better or the same as they were expecting^54^. 93% took analgesia This is consistent with mean maximum pain scale of 6-8 reported elsewhere^55^. In one study half of people experienced more bleeding than they anticipated^37^ and in another 77% of participants reported that it was more than their usual menstrual period^54^.

The pain relief offered to people completing medical abortion at home is not standard across services. When people completing a medical abortion at home were provided with a combination of paracetamol, ibuprofen and rectal diclofenac, almost all used all three medications, one quarter accessed additional analgesia and 65% used additional non-pharmacological measures such as heat and rest to reduce pain^37^. In recent UK studies, 93% used pain relief (41-92% paracetamol, 39-61% ibuprofen 64% dihydrocodeine/codein)^25,54^.

Most people who experienced significant pain and bleeding felt more comfortable coping with this at home, “…it is such a physical and emotional process so, y’know, at home’s better. You basically need to make friends with your toilet for 8 hours, so at home’s better. I was such a mess. I would be embarrassed for people to see me like that.”^38^. However, a minority would have preferred to have in-person health professional care because of the practical and emotional support provided^37,24,23^.

When people having medical abortion at home have had a previous pregnancy (ending in live birth, abortion or miscarriage), they are less likely to report pain that is distressing or to request professional support in comparison with those who have not had a previous pregnancy. However, they are no less likely to report heavy bleeding^37^. One reason to choose medical abortion at home is that people wish to be aware of the process of the abortion. Some find it helpful to know that the pregnancy has been passed if they see the products of conception, while others find it distressing^38^.

Some people having an abortion valued the support of partners, family and friends for company or practical support while others valued the opportunity to complete the abortion alone^24^. Partners (all men in cited study) interviewed about their experience of supporting people having a medical abortion expressed feelings of sadness, worry and concern, and sometimes ambivalence, about the decision to have an abortion as well as valuing the opportunity to support their partners through the process ^56^.

#### Assessing the success of abortion

There is good evidence to suggest that people can self-assess the success of abortion^57,58,59, 60,61,62^ (Table 5). There was little evidence in the literature of people’s thoughts and feelings about this process.

**Table 5:**
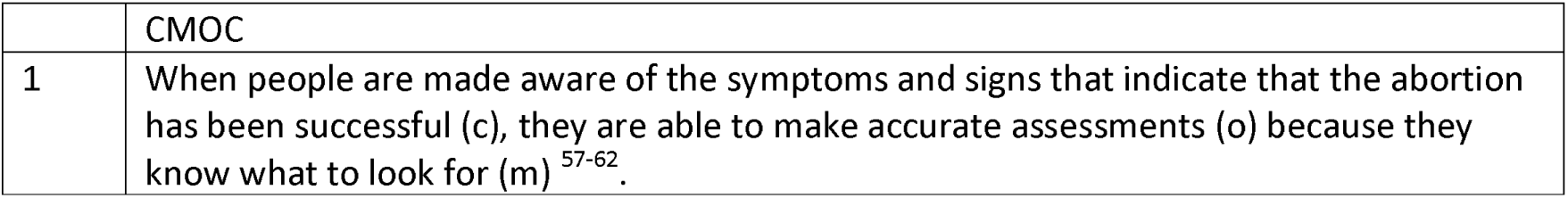
CMOC for ‘Assessing the success of abortion’

#### Choosing contraception

For some people the time of abortion provision is a convenient, appropriate and valued opportunity to discuss contraception and national/international guidelines recommend this^58^ (Table 6). For those people opting for implants, having them inserted when medical abortion medication is provided, or for those choosing an intrauetine devices, an appointment booked shortly after medical abortion, means they are more likely to receive their contraceptive of choice ^38,63,64^. However, medical abortion via telemedicine limits the opportunity to do this. For other people, a contraceptive discussion at the time of the medical abortion consultation is unwelcome^65^ and may be associated with a sense of being judged and pressurised about contraception^38^. Some people also prefer to focus on managing their abortion before considering contraception^38,65-67^. In this situation the offer of a delayed telephone or face to face contraception discussion can be welcome including for those who have had difficulty finding a contraceptive method that suits them^38,68,69,70,71^.

**Table 6:** CMOCs for ‘Choosing contraception’

|  | CMOC |
| --- | --- |
| 1 | At the time of abortion provision (c) some people are happy to discuss or be provided with contraception (o) because of the salience of this issue for them and the convenience (m) <sup>38</sup> |
| 2 | When people opting for long-acting reversible contraceptives (LARC) have an appointment booked |
|  | for them when receiving abortion medication at a time and place of their choosing after the abortion, (c ) they are more likely to receive their contraception of their choice (o) because of salience (m) and less work for users in making the appointment (m)<sup>63,64</sup> |
| 3 | When people who are focused on the abortion procedure are offered contraceptive advice at a later date (c), they are more likely to welcome a discussion about contraception (o), because they have the capacity to consider this (m)<sup>38, 68, 69, 70, 71</sup> |
| 4 | When people who have had a previous abortion are offered telephone contraceptive counselling following abortion (c) they are more likely to take this up (o) because they may want help with complex contraceptive needs (m)<sup>68, 70, 71</sup> |
| 5 | When contraception is raised at the time of abortion (c), people can feel judged (o) if health care providers imply that it is important to prevent future unintended pregnancies (m)<sup>38</sup> |
| C6 | When blended (digital linked to face to face) post abortion support and contraception services are designed with user input (c) they are likely to be acceptable and accessible to users (o) because they address their needs (m).<sup>72</sup> |

## Discussion

### Choice of abortion procedure remains essential

Increased expectations of self-care, access to digital health services and the COVID 19 pandemic may have reduced choice of abortion method. High levels of satisfaction are reported after medical abortion but variation in experience means that choice of method remains important^21,23,31,36^. Though medical abortion at home is a lower cost procedure than medical abortion in hospital which, in turn, is a lower cost procedure than surgical abortion^73^, personal preference, past experience or context may make some abortion processes, for some people, difficult, distressing or unsafe ^74,75^. People who may have lower levels of satisfaction with medical abortion at home include those who: have not had a previous pregnancy, do not have access to a comfortable and private space at home, do not have support from family/partners/friends, have inflexible personal or employment responsibilities, are homeless or wish to disengage from the process.

### Realistic information on the range of experience of abortion at home is important so that the abortion does not generate surprise or anxiety and so that people can adequately prepare for it at home

Informed decisions require information on the potential advantages and disadvantages of the available treatment options and abortion providers should offer information on the effectiveness, safety, health risks and benefits and the expected range of experiences of all abortion options. Although the effectiveness of abortion procedures is important to inform choice as up to 5% may require a surgical evacuation to complete the abortion^76-78^, effectiveness was not mentioned in user accounts of reasons for choosing medical abortion at home. Our review shows that it is important to communicate: a range of experiences, the importance of context (for example, where there is lack of privacy) or personal experience (for example, those with previous pregnancies report less pain). There is limited recent research on the best strategies to communicate this information and further work is required to collate and test good practice.

### Acknowledging the implications of moving a process from a health care setting to home

Guidelines on support for medical abortion at home may not sufficiently acknowledge the work that people do to prepare their space, manage their privacy, fulfil their work and caring obligations around the process, make decisions on pain relief, clean household areas, decide when and how to take misoprostol, assess their experience against what is normal and decide when to ask for professional help. There is a need to understand the new types of information/advice required to support people who do this work. The response may involve new technologies (e.g. medical abortion support at home apps), new or developing roles (e.g. medical abortion doulas) or new forms of peer support (e.g. discussion forums to help with preparation and share experience). Further research is required to understand and define health service responsibilities when procedures move from clinically facilitated to self-managed in general and for medical abortion at home in particular.

### Understanding optimal pain relief

There is no consensus within the literature on analgesia for those who experience pain during medical abortion although a Cochrane Review is currently in progress on this topic^79^. There is currently significant variation in practice described in terms of the analgesics offered, the schedule and route of administration and whether these are provided by health services.

### More choice within the procedure

Enabling people to tailor their abortion experience could be an empowering strategy within a process that is often experienced as disempowering^32,80^. Options include when and how to take medications, pharmaceutical and other strategies for pain relief, the amount and form of support available and when contraception is discussed.

### Contraception

Peri-abortion contraception counselling should be delivered without pressure to take up contraception as some people find this stigmatising. A choice of when and where to discuss contraception is important. The option of delayed counselling/provision is helpful for some and may be the only option for those who are having intrauterine devices or implants and choose telemedicine abortion.

#### Strengths and limitations

We were systematic and transparent in our approach to the realist review which was conducted in accordance with the RAMSES standards (27). Our authorship team represents a variety of clinical and academic backgrounds, ensuring divergence in our analysis and we benefited from expert feedback from the multi-disciplinary SACHA team throughout the review. Limitations include our analysis on publicly accessible literature, located through recognised research databases and Google. There were clear gaps in the evidence that we found and we have highlighted these in our conclusions.

#### Further research needed

Based on our review of the literature we recommend that further research is needed on:

- Optimal strategies for providing information and supporting choice of procedure/setting.
- Understanding how medical abortion at home involves or impacts family and friends (beyond partners).
- Optimal medications for pain relief including the dosing, route of administration and length of time that pain relief is required.
- How to offer choice within the abortion procedure in a manner that is empowering and not overwhelming
- Optimal means for providing contraception advice /support after the abortion procedure.

#### Recommendations

Our recommendations are summarized in our revised programme theory in figure 4 below.

**Figure 4:**
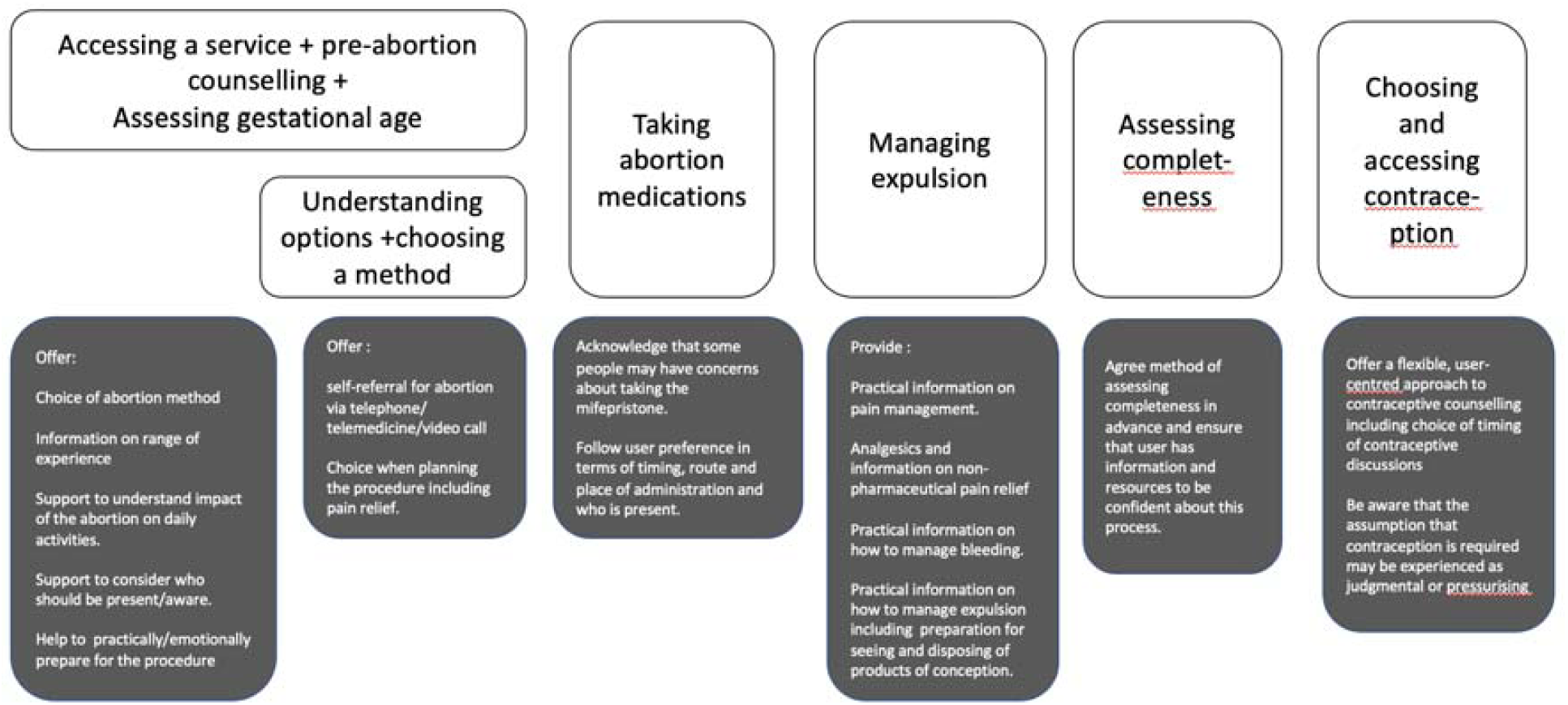
Revised programme theory with key learning from this review

- People should be given a choice of type and place of abortion
- Discussion of choice of method should include service factors, personal responsibilities, geographical context and preferred level of awareness of/engagement with the procedure
- Information to support choice of procedure should include the range of experience of medical abortion and realistic accounts of pain and bleeding.
- Health system support for the abortion process should acknowledge the new types of work that people take on when they manage this process at home, for example preparing a space for the abortion.
- There should be opportunities to personalize experience including when and where to take medications, who is present, where to pass products of conception.
- Support for taking mifepristone should acknowledge the emotional significance of this for some people.
- Adequate pain relief is an essential part of satisfaction with medical abortion, particularly for those who are pregnant for the first time. Pain relief should be offered as well as information on non-pharmacological strategies to manage pain.
- People should be offered a choice of where and when to discuss contraception including opportunities to book LARC appointments if this is their preference.

## Conclusions

Acknowledging the work done by patients when moving an intervention from clinic to home is important. This includes preparing a space, managing privacy, managing work/caring obligations, deciding when/how to take medications, understanding what is normal, assessing experience and deciding when and how to ask for help. Strategies that reduce surprise or anxiety and enable preparation and a sense of control support the transition of this complex intervention outside healthcare environments. It is important that options for those who are unable to take on these additional responsibilities or who prefer not to do so are maintained including clinical facilities and appropriately trained clinicians who can provide surgical abortion and medical abortion within health care facilities.

## Data Availability

Data extracted from studies and used for analysis can be requested from the corresponding authors.  

## Author contributions

RF, KW, CF & PB conceptualised the study. GW was responsible for the methodology. RF, KW, CF, PB, GW, RM, MP, RS and ML wrote the protocol. ML registered the review in the Prospero system (ID CRD42021225307) and conducted the searches. All authors screened the papers. WN conducted preliminary data extraction. All authors participated in the data analysis. AI composed the tables. PB, CF, WN and GW wrote the first draft. All authors contributed to the final manuscript. This paper reports on data collected as part of the SACHA Study: Shaping Abortion for Change, funded by the National Institute of Health Research. (www.lshtm.ac.uk/sacha) Collaborators of the SACHA Study provided feedback on all stages of the study.

## Protocol

Protocol can be found in Propsero under <u>ID CRD42021225307</u>

## Data availability statement

Data extracted from studies and used for analysis can be requested from the corresponding authors.

## Appendix 1: Databases searched and search terms

Ovid MEDLINE(R) and Epub Ahead of Print, In-Process & Other Non-Indexed Citations, Daily and Versions(R) 1946 to December 8, 2021; Embase Classic+Embase 1947 to 2021 December 08; APA PsycInfo 1806 to November Week 5 2021; APA PsycExtra 1908 to November 08, 2021; Global Health 1910 to 2021 Week 49; Social Policy and Practice 202010; CINAHL Plus, Web fo Science Core collection; Scopus (Elsevier); Cochrane Library.

Seach terms for Ovid: ((exp Abortion, Induced/) OR (abortion) OR (abortions) OR (pregnancy adj4 terminat*)) AND ((self care/) OR (self-management/) OR (Self Administration/) OR (Telemedicine/) OR (exp communications media/) OR (Internet-Based Intervention/) OR (Text Messaging/) OR (Outpatients/) OR (nurses/) OR (nursing staff/) OR (Nurse Midwives/) OR (Midwifery/) OR (Pharmacists/) OR (exp General Practice/) OR (Primary Health Care/) OR (Patient-Centred Care/) OR (self-manag*) OR (self-administer*) OR (self-induc*) OR (self-assess*) OR (self-sourc*) OR (“at home”) OR (“home use”) OR (home adj4 dosage) OR (home adj4 administ*) OR (home adj4 abortion) OR (telemedic*) OR (digit*) OR (online) OR (web) OR (website) OR (internet) OR (mobile) OR (sms) OR (text*) OR (messag*) OR (app) OR (“social media”) OR (outpatient) OR (task-shar*) OR (task-shift*) OR (mid-level) OR (midlevel) OR (nurs*) OR (midwi*) OR (pharmacist) OR (pharmacy) OR (GP) OR (GPs) OR (general adj3 practic*) OR (non-speciali*) OR (non-clinical) OR (licen*) OR (qualif*) OR (decrim*) OR (woman-centered) OR (woman-centred) OR (women-centered) OR (women-centred) OR (patient-centered) OR (patient-centred) OR (gestation* adj5 limit*)) AND ((“Patient Acceptance of Health Care”/) OR (Health Services Accessibility/) OR (exp patient satisfaction/ or patient preference/) OR (access*) OR (safe*) OR (accept*) OR (complicat*) OR (side effect*) OR (efficac*) OR (effect*) OR (feasib*) OR (prefer*) OR (satisf*) OR (feel*) or (autonom*) OR (delay*) OR (wait*) OR (experienc*) OR (uptake*) OR (time*) OR (distan*) OR (succe*) OR (impact) OR (opinion*) OR (best practice*) OR (capacity) OR (capab*) OR (competen*))

We searched Google (limited to the first 100 results) to identify grey literature and included known documents suggested by experts within the team and those identified through hand searches of the references of retrieved articles. Results from 01/01/2000 and 09/12/2022 that were peer reviewed research (journal articles and conference proceedings) theses or grey literature were included. We included papers where only a part of the investigation was relevant. We did not include books, book sections, cases, serials.

## Appendix 2: Questions used to identify relevance

Five questions to operationalise a realist logic of analysis, adapted from Duddy et al (2021).

- Is there content that provides information on the context, mechanism or outcome of interventions? (CMO value)
- What do these data tell us about the context, mechanism or outcome of the interventions studies? (CMO outcomes)
- How should the CMO outcomes enable refinement of our initial programme theory? (refinement)

## Appendix 3: Summary of papers included in the review

| First Author | Date | Title | Source Type | Country | Setting | Participant Type | Number of Participants | Study Design | Is the paper relevant to the question? | Are the methods used appropriate? | Are the findings plausible? | Do conclusions support those of other studies? |
| --- | --- | --- | --- | --- | --- | --- | --- | --- | --- | --- | --- | --- |
| Aiken, A. R. A | 2018 | Motivations and Experiences of People Seeking Medication Abortion Online in the United States | Original research paper | United States | Online | People seeking online medical abortion services | 32 | Qualitative interview study | Yes - paper explores motivations for considering self-managed abortion. | Yes - in-depth interviews were used to gather qualitative data on people's motivations and experiences of seeking online medical abortion. | Yes | Yes |
| Aiken, A. | 2021b | Demand for self-managed online telemedicine abortion in eight European countries during the COVID-19 pandemic: a regression discontinuity analysis | Original research paper | Germany; Hungary; Italy; Malta; Netherlands; Northern Ireland; Portugal; Great Britain | Online | N/A | 3915 requests for self-managed abortion to Women on Web (WoW) | Cross-sectional comparative analysis of requests for self-managed abortion pre- and post the Covid-19 pandemic | Yes - paper assesses whether the COVID-19 pandemic increased demand for self-managed medical abortion provided through online services and suggests reasons for trends in demand for online accessed abortion during the pandemic. | Yes - request rates in 8 countries are compared using regression discontinuity both before and after lockdown measures. | Yes | Yes |
| Atay | 2021 | Why women choose at-home abortion via teleconsultation in France: drivers of telemedicine abortion during and beyond the COVID-19 pandemic. | Original research paper | France | Online | People accessing online medical abortion services | 140 | Mixed Methods (Cross-sectional survey of consultation data and email content analysis) | Highly - the paper explores drivers of at home, online supported, medical abortion during the pandemic and beyond. | Yes - the findings are based on appropriate analysis of consultations and emails extracted from an online medical abortion service to reveal the motivations and preferences of women seeking at home abortion. | Yes | Yes |
| Boydell | 2021 | Women's experiences of | Original research | Scotland | Online | People accessing | 20 | Qualitative interview study | Highly - paper explores women's | Yes - the findings are based on appropriate use of | Yes | Yes |
|  |  | a telemedicine abortion service (up to 12 weeks) implemented during the coronavirus (COVID-19) pandemic: a qualitative evaluation | paper |  |  | online medical abortion services |  |  | experiences of accessing online medical abortion services. | qualitative interviews to explore women's experiences of telephone consultation; remote support; views on no pre-abortion ultrasound; and self-administration of abortion medications at home. |  |  |
| Ehrenreich | 2019 | Spatial dimensions of telemedicine and abortion access: a qualitative study of women's experiences | Original research paper | Utah, USA | Online | People accessing online medical abortion services | 20 | Qualitative interview study | Yes - the paper explores women's experiences of using online services for the first mandatory step when accessing medical abortion in Utah, an 'information visit'. | Yes - through in-depth interviews thoroughly explore experiences of using online services for information visits to fulfil abortion requirements. | Yes | Yes |
| Endler | 2019 | Telemedicine for medical abortion: a systematic review | Review | N/A | N/A | N/A | N/A | Systematic Review | Yes - the paper assesses the success rate, safety, and acceptability for women and providers of online medical abortion services. | Yes - a systematic review assessing a broad and significant number of relevant studies. | Yes | Yes |
| Erlank | 2021 | Acceptability of no-test medical abortion provided via telemedicine during Covid-19: analysis of patient-reported outcomes | Original research paper | United Kingdom | Online | People accessing online medical abortion services | 1243 | Qualitative survey study | Yes - the study reports patient-reported outcome measures assessing the quality of consultation, access to medicine after consultation, ability to manage the process at home and overall patient satisfaction. | Yes – early medical abortion patients invited to opt-in to a follow-up call post-procedure to answer clinical and satisfaction questions to measure acceptability of the service. | Yes | Yes |
| Eshre Capri Workshop Group | 2017 | Induced abortion | Workshop | N/A | N/A | N/A | N/A | N/A | Moderately - This paper presents commentary on induced abortion gathered at a workshop of experts in the field. The commentary included discussion of at home medical abortion with reference to online support. | N/A | N/A | Yes |
| Finch | 2019 | Impact of self-administration of misoprostol for early medical abortion: a prospective cohort study | Original research paper | Scotland | Home | People accessing abortion services both at home and at a facility | 2430 | Prospective observational study | Yes - the study explores the impact of Scotland's legalisation on home use of misoprostol for the purpose of early medical abortion on uptake and success rate, and on the provision of effective contraception on discharge. | Yes - prospective observational study followed the outcomes of two cohorts who received their abortion care, before and after the introduction of home use of misoprostol. | Yes | Yes |
| Fix | 2020 | At-home telemedicine for medical abortion in Australia: a qualitative study of patient experiences and recommendations | Original research paper | Australia | Online | People accessing online medical abortion services | 24 | Qualitative interview study | Highly - the study explores people's experiences obtaining a medical abortion through an online service and discusses people's reasons for choosing an online service. | Yes - in-depth interviews appropriately captures the experiences of people using the at-home telemedicine service. | Yes. | Yes |
| Gill | 2019 | Feasibility and Acceptability of a Mobile Technology Intervention to Support Postabortion Care in British Columbia: Phase 1 | Original research paper | Canada | Abortion care facility | People attending an abortion care facility | 58 | Mixed-methods formative study (cross-sectional survey and semi-structured interviews) | Moderately - study aims to understand how people at surgical abortion clinics utilise their mobile phones to access health care information and their preferences for a mobile intervention that supports follow-up care. | Yes - mixed-methods study appropriately explores a variety of aspects relevant to technology usage during abortion process. | Yes | Yes |
| Gill & Norman | 2018 | Telemedicine and medical abortion: dispelling safety myths, with facts | Editorial | N/A | N/A | N/A | N/A | Editorial commentary on recent research regarding online medical abortion services | Moderately - This paper presents commentary on recent research regarding online medical abortion services. | N/A | N/A | Yes |
| Goldman | 2021 | Transcutaneous Electrical Nerve Stimulation to Reduce Pain With Medication Abortion: A Randomized Controlled Trial | Original research paper | N/A | Abortion care facility | People undergoing medical abortion. | 251 | Randomised controlled trial (RCT) | Yes - evaluates whether high-frequency transcutaneous electrical nerve stimulation reduces experiences of pain during medical abortion. | Yes – randomized control trial shows that participants who received high-frequency transcutaneous electrical nerve stimulation after medical abortion had significantly lower posttreatment pain scored compared to those that received a sham procedure. | Yes | Yes |
| Hamoda | 2005 | The acceptability of home medical abortion to women in UK settings | Original research paper | England and Scotland | Home | People seeking hospital based abortion care | 553 | Qualitative survey study | Highly - the study explores the acceptability of home medical abortion to people UK settings including managing pain and bleeding at home. | Yes - self-complete questionnaires adequately measure people's views on the acceptability of medical abortion at home. | Yes | Yes |
| Harden | 2021 | Women's experiences of self-administration of misoprostol at home as part of early medical abortion: a qualitative evaluation | Original research paper | Scotland | Home | People that had recently undergone early medical abortion | 20 | Qualitative interview study | Highly - the study explores the experiences of people who accessed at home medical abortion and reported benefits of at home administration including flexibility, privacy and comfort. | Yes - qualitative data is collected exploring what people desire and valued during the process. | Yes | Yes |
| Heath | 2019 | A comparison of termination of pregnancy procedures: Patient choice, emotional impact and satisfaction with care | Original research paper | Sweden | Abortion care facility | People requesting an abortion |  | A mixed-method prospective comparative study | Yes | Yes | Yes | Yes |
| Hedqvist | 2016 | Women's experiences of having an early medical abortion at home | Original research paper | Sweden | Home | People accessing medical abortion at home | 119 | Cross-sectional (semi-structured telephone interviews) | Highly - paper investigates people's experiences of having a medical abortion at home including experiences of pain and bleeding. | Yes - descriptive and comparative design enable an in-depth and balanced conclusion. Study also investigates differences between groups of people. | Yes | Yes |
| Hoggart and Berer | 2021 | Making the case for | Editorial | N/A | Online | N/A | N/A | Editorial commentary on | Moderately – this paper reviews recent | N/A | N/A | Yes |
|  |  | supported self-managed medical abortion as an option for the future |  |  |  |  |  | recent research regarding online medical abortion services | research on online medical abortion services highlighting benefits and pointing out need for further development of services, suggesting a need for access to 24 hours support. |  |  |  |
| Iyer | 2021 | Preferences for contraceptive counselling and access among abortion patients at an independent clinic in Texas | Original research paper | USA | Abortion care facility | People attending an abortion care facility | 181 | Cross-sectional survey study | Yes - study explores preferences for contraceptive counselling and access to contraception. | Yes - self-administered surveys used as an appropriate method considering legally restrictive setting and associated stigma. | Yes | Yes |
| Kavanagh | 2011 | Patients' attitudes and experiences related to receiving contraception during abortion care | Original research paper | USA | Abortion care facility | People accessing abortion at an abortion care facility | 542 | Cross-sectional survey study | Yes - study documents attitudes towards contraceptive services and identifies participants characteristics associated with desire for contraception and interest in LARC. | Yes - appropriate methods for exploring attitudes and characteristics associated with specific needs relevant to contraceptive use. | Yes | Yes |
| Kero | 2010 | Home abortion - experiences of male involvement | Original research paper | Sweden | Home | Couples - where the male partner had been present when their female partner had had an at home medical abortion | 23 couples | Qualitative interview study | Moderately - the paper explores the male partner's experience of being present and supporting their partner during an induced home abortion. | Yes - qualitative method ensures high detailed analysis in experiences of both the pregnant woman and her partner. | Yes | Yes |
| Killinger | 2022 | Why women choose abortion through telemedicine outside the formal health sector in Germany: a mixed-methods study | Original research paper | Germany | Online | People accessing an online medical abortion service | 1090 | Cross-sectional study analysing data from online consultations | Yes - the paper aims to understand the motivations and barriers to access for people who choose online abortion services outside of the formal health sector. | Yes - a cross-sectional study of data contained in online consultations and a content analysis of over 100 email texts. | Yes | Yes |
| Levine and Cameron | 2009 | Women's preferences for method of abortion and management of miscarriage | Original research paper | United Kingdom | Abortion care facility | People undergoing medical abortion, surgical abortion and surgical management of miscarriage | 148 | Cross-sectional survey study | Yes - the study explores views on medical treatment at home, and surgery under local anaesthesia, to determine whether new services should be developed. | Moderately - small sample size of self-administered anonymous questionnaires. | Yes | Yes |
| Loeber | 2016 | Contraceptive counselling for women with multiple unintended pregnancies: the abortion client's perspective | Original research paper | Netherlands | Abortion care facility | Survey – People attending an abortion care facility<br>Interviews – People attending an abortion care facility that had had more than three unintended pregnancies | 212 | Mixed methods (quantitative survey and semi-structured interviews) | Yes - study explores views on contraception use after induced abortion and post-abortion contraceptive counselling | Yes - mixed method approach appropriately explores experiences of post-abortion contraception and contraception counselling both in-depth and across a larger sample. | Yes | Yes |
| Lohr | 2018 | Telephone or integrated contraception counselling before abortion: impact on method choice and receipt | Original research paper | United Kingdom | Abortion care facility | People who had accessed abortion at an abortion care facility | 18,573 | Cross-sectional study of de-identified data | Yes - paper compares the characteristics of people who chose contraception counselling either over the telephone and separate from abortion consultation or face-to-face and integrated into consultation. | Yes - very large sample size which thoroughly explores demographic characteristics and contraceptive method choice. | Yes | Yes |
| Lokeland | 2014 | Medical abortion with mifepristone and home administration of misoprostol up to 63 days' gestation | Original research paper | Norway | Abortion care facility | People seeking an at home medical abortion before 63 days' | 1,018 | Observational study | Yes - paper evaluates the acceptability of medical abortion at home, including pain and bleeding. | Yes – a large sample size of people experiencing at home abortion with follow up telephone recorded to assess bleeding, pain and acceptability. | Yes | Yes |
| Low | 2021 | Women's experiences of self-referral to an abortion service: | Original research paper | Scotland | Abortion care service | People attending an abortion care service | 21 | Qualitative interview study | Yes - this study evaluates a self-referral service to abortion services by investigating its | Yes - semi-structured interviews to focus on people's experiences of the referral process. | Yes | Yes |
|  |  | qualitative study |  |  |  |  |  |  | impact on people's experiences of the referral process. |  |  |  |
| Makenzius | 2012 | Autonomy and dependence - experiences of home abortion, contraception and prevention | Original research paper | Sweden | Home | People who have experienced an at-home abortion | 37 | Qualitative interview study | Yes - study explores people's experiences and needs related to care in the context of a home abortion. | Yes – interviews capture patients at-home abortion experiences and needs which fall under themes of autonomy (choice of at-home abortion, increase of privacy and control) and dependence (desire to be treated with empathy by health care workers and receive adequate information). | Yes | Yes |
| Matulich | 2014 | Understanding women's desires for contraceptive counselling at the time of first-trimester surgical abortion | Original research paper | USA | Abortion care facility | People receiving surgical abortion at an abortion care facility | 199 | Qualitative survey study | Yes - paper investigates whether or not people presenting for a first-trimester surgical abortion want to discuss contraception on the day of their procedure. | Yes - survey obtained demographic information and inquired about desire for contraceptive counselling. | Yes | Yes |
| Norman | 2014 | Access to Complex Abortion Care Service and Planning Improved through a Toll-Free Telephone Resource Line | Original research paper | Canada | Abortion care facility | N/A | N/A | Review of service provision model | Yes - study presents a review of a service provision model which has provided improved access to abortion care through a toll-free telephone abortion access and counselling service. | Yes - a descriptive analysis appropriately reports on the development and delivery of this service. | Yes | Yes |
| Oppegaard | 2015 | Clinical follow-up compared with self-assessment of outcome after medical abortion: a multicentre, non-inferiority, randomised, controlled trial | Original research paper | Austria; Finland; Norway; Sweden | Abortion care services | People requesting medical abortion | 924 | Randomised, controlled, non-inferiority trial | Yes - study compares clinical assessment with self-assessment of abortion outcome. | Yes - assigned people in a 1:1 ratio to attend routine clinical follow-up or to self-assess outcome at home with a semi-quantitative urine human chorionic gonadotropin test 1-3 weeks after abortion. | Yes | Yes |
| Pohjoranta | 2018 | Predicting poor | Original research | Finland | Abortion care | People undergoing a | 605 | RCT | Moderately - paper assesses factors | Yes – the study compared the intervention group | Yes | Yes |
|  |  | compliance with follow-up and intrauterine contraception services after medical termination of pregnancy | paper |  | facility | medical abortion |  |  | associated with non-compliance with post-abortion services and evaluates differences in rates of attendance and intrauterine device insertion. | booked to have IUD insertion 1-4 weeks after medical abortion and women in control group were advised to contact their PHC for follow-up and IUD insertion. |  |  |
| Pohjoranta | 2020 | Early provision of intrauterine contraception as part of abortion care - 5-year results of a randomised controlled trial | Original research paper | Finland | Abortion care facility | People undergoing an induced abortion | 748 | RCT | Moderately - paper investigates whether the incidence of subsequent termination of pregnancy can be reduced by providing IUD as part of the abortion service. | Yes – RCT assesses the effectiveness of early comprehensive provision of IUD after induced abortion. Intervention group provided with an IUD during surgical abortion and women in control group advised to contact primary healthcare for follow-up and IUD insertion. | Yes | Yes |
| Powell-Jackson | 2010 | Benefits of using a digital video disk for providing information about abortion to women requesting termination of pregnancy | Original research paper | Scotland | Abortion care facility | People attending an abortion care facility and facility staff | 226 | Comparative cross-sectional survey design | Yes - paper examines the benefits of using a DVD for providing information about abortion. | Yes - questionnaires are used to appropriately measure people's satisfaction with the information received through the DVD and what more/less they would want it to include. | Yes | Yes |
| Purcell | 2017 | Self-managment of first trimester medical termination of pregnancy: a qualitative study of women's experiences | Original research paper | Scotland | Abortion care facility | People who are administered a medical abortion at a facility and return home to complete termination | 44 | Qualitative interview study | Yes - explores people's experiences of returning home to complete medical abortion, including self-monitoring of treatment process. | Yes - in-depth interviews to assess people's experiences in detail. | Yes | Yes |
| Rafferty & Longbons | 2021 | #AbortionChanges You: A Case Study to Understand the Communicative Tensions in Women's | Original research paper | United States | Online | People who have had a medical abortion | 98 blog posts | Case study | Yes - study analyses people's narratives after having had a medication abortion though blogs posted on a website - Abortion Changes | Yes - using a case study approach enabled the study to explore multiple perspectives rooted in specific contexts. | Yes | Yes |
|  |  | Medication Abortion Narratives |  |  |  |  |  |  | You. |  |  |  |
| Raymond | 2018 | Low-sensitivity urine pregnancy testing to assess medical abortion outcome: A systematic review | Review | N/A | N/A | N/A | N/A | Systematic review | Yes - the study explores data on the accuracy and acceptability of a strategy for identifying ongoing pregnancy after medical abortion treatment using a low-sensitivity pregnancy test (LSPT). | Moderately - A more comprehensive search across other databases could strengthen study. | Yes | Yes |
| Raymond | 2017 | Self-assessment of medical abortion outcome using symptoms and home pregnancy testing | Original research paper | United States | Abortion care facility | People undergoing medical abortion | 343 | Intervention study | Highly - study evaluates compliance with a strategy to enable medical abortion patients to assess treatment outcome on their own and decide whether to seek clinical follow-up. | Yes - appropriate methodology to clinically assess women's compliance to using a pregnancy test after medical abortion and women's ability to self-assess treatment outcome. | Yes | Yes |
| Reynolds-Wright | 2020 | Pain management for medical abortion before 14 weeks' gestation | Review | N/A | N/A | N/A | N/A | Systematic review | Yes - paper reviews pain relief regimens for the management of medical abortion. | Yes - wide variety of databases, conference abstracts and organisations systematically searched and analysed. | Yes | No |
| Reynolds-Wright | 2021 | Telemedicine medical abortion at home under 12 weeks' gestation: a prospective observational cohort study during the COVID-19 pandemic | Original research paper | Scotland | Online/Home | People choosing online medical abortion services | 663 | Prospective cohort study | Yes - the paper determines the efficacy of an online at-home medical abortion service, noting any complications after treatment and acceptability of care. | Yes - large sample size. Questionnaires and hospital database analyses allows for in-depth review of efficacy, complications and acceptability. | Yes | Yes |
| Robson | 2009 | Randomised preference trial of medical versus surgical termination of pregnancy less than 14 weeks' | Original research paper | United Kingdom | Abortion care facility | People undergoing induced abortion and people attending a contraceptive | 1,877 | Randomised preference trial and economic evaluation with qualitative sub study | Yes - the paper explores the acceptability, efficacy and costs of medical abortion compared with surgical abortion and aims to understand | Yes – participants are either randomly assigned a procedure type (with consent to random allocation) or choose their procedure type to evaluate whether in people without | Yes | Moderately |
|  |  | gestation (TOPS) |  |  |  | and sexual health clinic |  |  | people's decision-making processes and experiences. | prior preference, acceptability of medical and surgical abortion is the same. A qualitative sub study explores people's preferences and decision making. |  |  |
| Rowlands & Wale | 2020 | A Constructivist Vision of the First-Trimester Abortion Experience | Constructivist Vision | N/A | N/A | N/A | N/A | Constructivist vision commentary on how a high-quality abortion experience might be achieved in the first trimester if regulatory laws were dismissed. | Moderately - This paper provides commentary on recent research on induced abortion in the first trimester within the framework of a future vision of state obligation to provide high quality abortion care. This paper also provides an evidence-based set of ingredients that facilitate a positive high-quality abortion experience. | N/A | N/A | Yes |
| Schmidt-Hansen | 2020 | Follow-up strategies to confirm the success of medical abortion of pregnancies up to 10 weeks' gestation: a systematic review with meta-analyses | Review | N/A | N/A | N/A | N/A | Systematic review | Yes - the review compares the effectiveness, safety, and acceptability of in-clinic and remote/self-assessment for confirming the success of medical abortion. | Yes - systematic review including RCT's from 2000 onward to ensure a broad range of studies captured. Quality of evidence compromised by small event rates, lack of blinding, and high attrition rates. | Yes | Yes |
| Sherman | 2017 | Providing experiential information on early medical abortion: a qualitative evaluation of an animated personal account, *Lara's Story* | Original research paper | Scotland | Abortion care facility | People who have had a medical abortion | 13 | Qualitative interview study | Yes - the study evaluates the views on an animated film, and its potential usefulness in providing information on medical abortion. | Yes - the use of interviews allows people to report their full experience and opinion in detail, also providing feedback or criticism. | Yes | Yes |
| Smith | 2019 | Current barriers, facilitators and future improvements to advance quality of abortion care: views of women | Original research paper | Edinburgh, Scotland | Abortion care facility | People seeking induced abortion | 154 | Cross-sectional survey study | Yes - the study explores current barriers access to care, and what future improvements should be implemented to abortion care services | Yes - self-administered anonymous questionnaires to capture experiences. | Yes | Yes |
| Smith | 2017a | Women's views and experiences of a mobile phone-based intervention to support post-abortion contraception in Cambodia | Original research paper | Cambodia | Abortion care facility | People who had received a mobile phone based post-abortion contraception counselling service. | 15 | Qualitative interview study | Yes - the study assesses views and experiences of receiving the MOBILE Technology for Improved Family Planning (MOTIF) intervention, which aims to increase post-abortion contraception. | Yes - semi-structured interviews to capture in-depth data. Small sample size may be a limitation. | Yes | Yes |
| Smith | 2017b | Process evaluation of a mobile phone-based intervention to support post-abortion contraception in Cambodia | Original research paper | Cambodia | Abortion care facility | People who had received a mobile phone based post-abortion contraception counselling service. | 249 | Process evaluation | Yes - the study assesses participants' interaction with the MOTIF intervention. | Yes - thorough process evaluation to assess associations with the intervention and post-abortion contraception use. | Yes | Yes |
| Upadhyay, U. D. | 2021 | Safety and Efficacy of Telehealth Medication Abortions in the US During the COVID-19 Pandemic | Original research paper | USA | Online | People accessing online medical abortion | 141 | Retrospective cohort study | Yes - the study assesses the safety and efficacy outcomes of a online medical abortion model. | Yes – appropriate methodology for assessing an online medical abortion service however the small sample size with some loss to follow-up, and thus some adverse events and ongoing pregnancies may have been undetected. | Yes | Yes |
| Whitehouse | 2021 | *It's a small bit of advice, but actually on the day, made such a difference....*: perceptions of quality in abortion care in England and | Original research paper | England; Wales | Abortion care facility | People who have had an abortion at a facility withing the last 6 months | 24 | Qualitative interview study | Yes - the paper explores participants' experiences and views on abortion quality of care. | Yes - interviews are an appropriate methodology to assess experiences. | Yes | Yes |
|  |  | Wales |  |  |  |  |  |  |  |  |  |  |
| Wieße | 2008 | Access to Abortion: What Women Want From Abortion Services | Original research paper | Canada | Abortion care facility | People seeking an abortion | 441 | Qualitative survey and interview study | Moderately - the paper identifies barriers to access for women seeking induced abortion at a facility. | Yes - questionnaire accurately explores demographic information, perceived barriers to access, and what women desire from abortion services. Interviews captured in-depth analysis to access questions. | Yes | Yes |

## Footnotes

1 Page MJ, McKenzie JE, Bossuyt PM, Boutron I, Hoffmann TC, Mulrow CD, Shamseer L, Tetzlaff JM, Akl EA, Brennan SE, Chou R, Glanville J, Grimshaw JM, Hróbjartsson A, Lalu MM, Li T, Loder EW, Mayo-Wilson E, McDonald S, McGuinness LA, Stewart LA, Thomas J, Tricco AC, Welch VA, Whiting P, Moher D. The PRISMA 2020 statement: An updated guideline for reporting systematic reviews.

